# Heterogeneity of Irritability: Psychometrics of The Irritability and Dysregulation of Emotion Scale (TIDES-13)

**DOI:** 10.1101/2022.02.08.22270624

**Authors:** Andrew S. Dissanayake, Annie Dupuis, Paul D. Arnold, Christie Burton, Jennifer Crosbie, Russell J. Schachar, Tomer Levy

## Abstract

**Objective:** We designed The Irritability and Dysregulation of Emotion Scale (TIDES-13) to test whether irritability consisted of several sub-dimensions that would correlate differentially with internalizing/externalizing psychopathology, age, and gender.

**Method:** Parent-report (n = 3935, mean age = 8.9) and youth self-report (n = 579, mean age = 15.1) versions of TIDES-13 were administered to a population-based sample. Exploratory and confirmatory factor analyses were conducted on separate sub-samples. We fit multivariable regression models between TIDES-13 sub-dimensions and age, gender, anxiety, depression, ODD and ADHD trait levels.

**Results:** A higher order model with a global irritability dimension and Proneness to Anger, Internalized Negative Emotional Reactivity, Externalized Negative Emotional Reactivity and Reactive Aggression sub-dimensions showed good to excellent fit in both parent-report and self-report. The global irritability dimension had a strong influence on all item variance (ω_Hierarchical; parent report_ = .0.94, ω_Hierarchical; self report_ = .90). Proneness to Anger, Externalized Negative Emotional Reactivity and Reactive Aggression decreased with age in males, whereas Internal Negative Emotional Reactivity increased with age in females. Internalized Negative Emotional Reactivity was associated with internalizing traits, over and above global irritability. ODD and ADHD were predicted primarily by the global irritability.

**Conclusion:** Although irritability can be estimated as an essentially unidimensional construct, differential associations of Internalized Negative Emotional Reactivity with gender and age, and internalizing psychopathology warrant examination in clinical populations. These results support TIDES-13 as a reliable and valid multidimensional measure of irritability and thus may be useful for research and clinical purposes.

## INTRODUCTION

Irritability is the one of the most common reasons for psychiatric evaluation and treatment in children and youth.^1,2^ Irritability is associated with a wide range of internalizing (eg. anxiety and depression)^3^ and externalizing psychopathology (eg. attention-deficit/hyperactivity disorder (ADHD)^4^ and oppositional defiant disorder (ODD^5^) and predicts significant concurrent and future impairment (eg. academic problems, poverty, and suicidality).^6,7^ Despite its importance, there is a lack of consensus on how to best conceptualize and measure irritability.^8^ This difficulty arises because irritability may manifest through a wide range of emotions, behaviors and internal states.^9^ For example, irritability may encompass excessive reaction to external (e.g., auditory stimulus, external cognitive distraction) or internal stimuli (e.g., fatigue or hunger) with negative emotions (ie. negative emotional reactivity),^10–12^ proneness to anger outbursts (i.e., “phasic irritability”), persistent irritable moods (i.e, tonic irritability),^13^ low tolerance for frustration in the face of blocked goal attainment (i.e., frustration intolerance),^14,15^ and a low threshold for maladaptive responses to threat with reactive aggression.^11,16^

Existing measures focus on a subset of possible manifestations of irritability, treating the construct as a homogeneous or unidimensional construct rather than a heterogeneous or multidimensional concept obscuring the possibility that sub-dimensions of irritability may exist and have distinct correlates. For example, the Affective Reactivity Index (ARI), a widely-used six item irritability scale in children and youth, primarily includes items that center on expressions of anger^17^ (ie.”stays angry for a long time”, “is angry most of the time”, “gets angry frequently”). On the other hand, the Bite Scale includes only items pertaining to an internal enduring state, alternatively characterized as tonic irritability^18,19^ (ie.”I have been grumpy, other people have been getting on my nerves”, “Things have been bothering me more than they normally do”, “I have been feeling irritable”). Despite these unidimensional conceptualizations, there is growing evidence for the hypothesis that irritability is a multidimensional trait with sub-dimensions that represent different states, emotions and behaviors.^19–22^ Thus, a measure that encompasses the variable sub-dimensions of irritability with strong internal reliability and discriminant validity would be an asset to clinical practice and scientific research progress.^8,9,17^

Prior to this study, we developed The Irritability and Dysregulation of Emotion Scale (TIDES-13) to measure a wide range of theoretical manifestations of the construct including proneness to anger (i.e., tonic irritability),^16^ anger outbursts (i.e., phasic irritability),^10 12,18^ negative emotional reactivity,^12^ and frustration intolerance^13^ (see supplementary materials). This present study assessed whether TIDES-13 captured a unified construct corresponding to irritability, as described previously in the literature, or whether irritability consists of sub-dimensions with distinct correlates. Using data from a large community sample, we used exploratory and confirmatory factor analyses to determine whether multiple irritability sub-dimensions could be discerned. Finally, we assessed the predictive validity of irritability sub-dimensions by examining their differential associations with age, gender, internalizing psychopathology and externalizing psychopathology.^19–22^ We also tested the hypothesis that a sub-dimension pertaining to anger would show the strongest levels of convergence with the ARI, given that the ARI mainly measures proneness to anger.^8^

## METHOD

Methods for Scale Development are described in the supplement.

### Participants

Visitors aged 4-19 years to a local science center (N = 4413) participated in this study in 2019. Sample characteristics are presented in Table 1. Parents were the informants for participants ages 4 to 19 (N = 3880); participants aged 10-19 (N = 533) provided self-report.

**Table 1:**
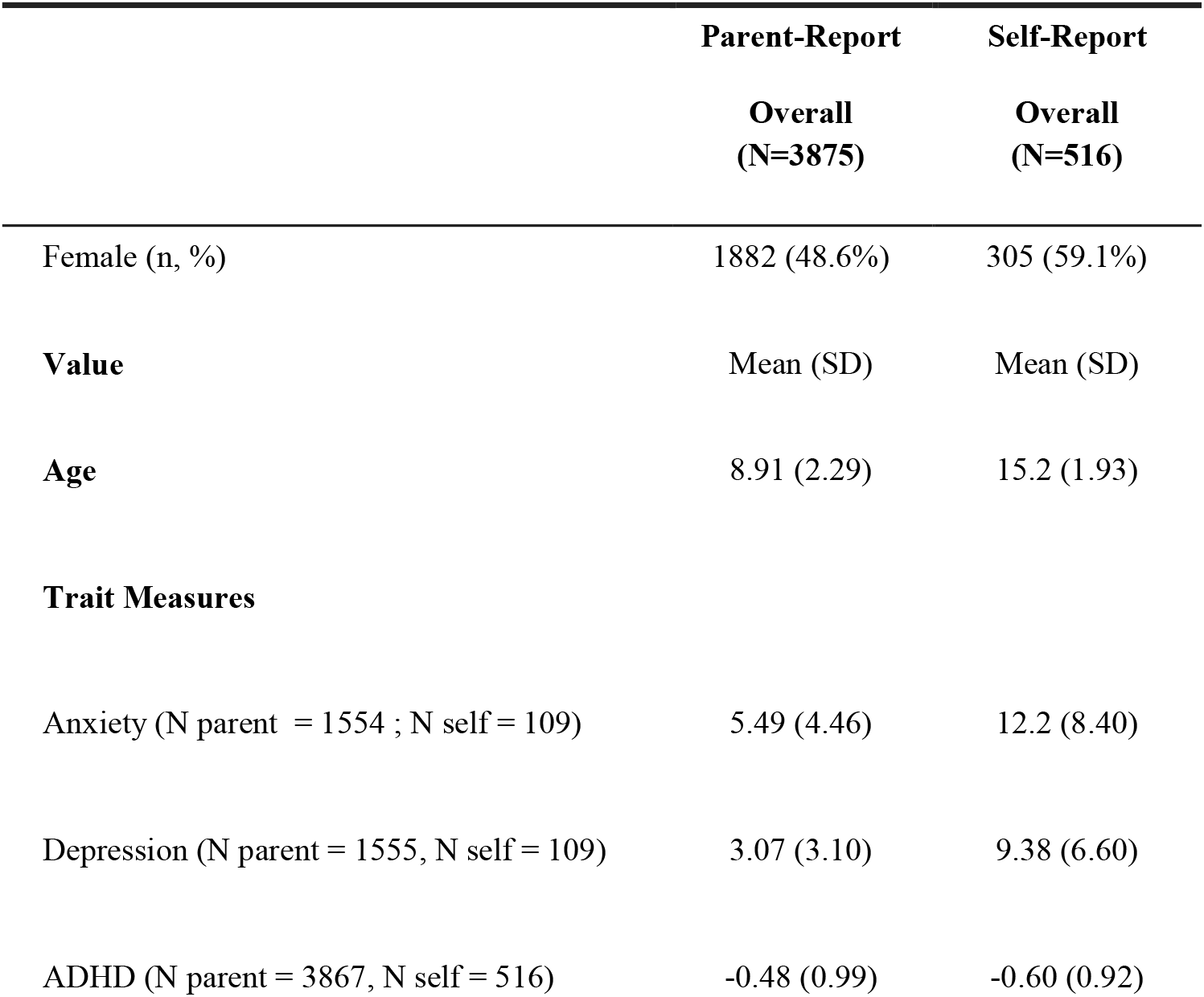
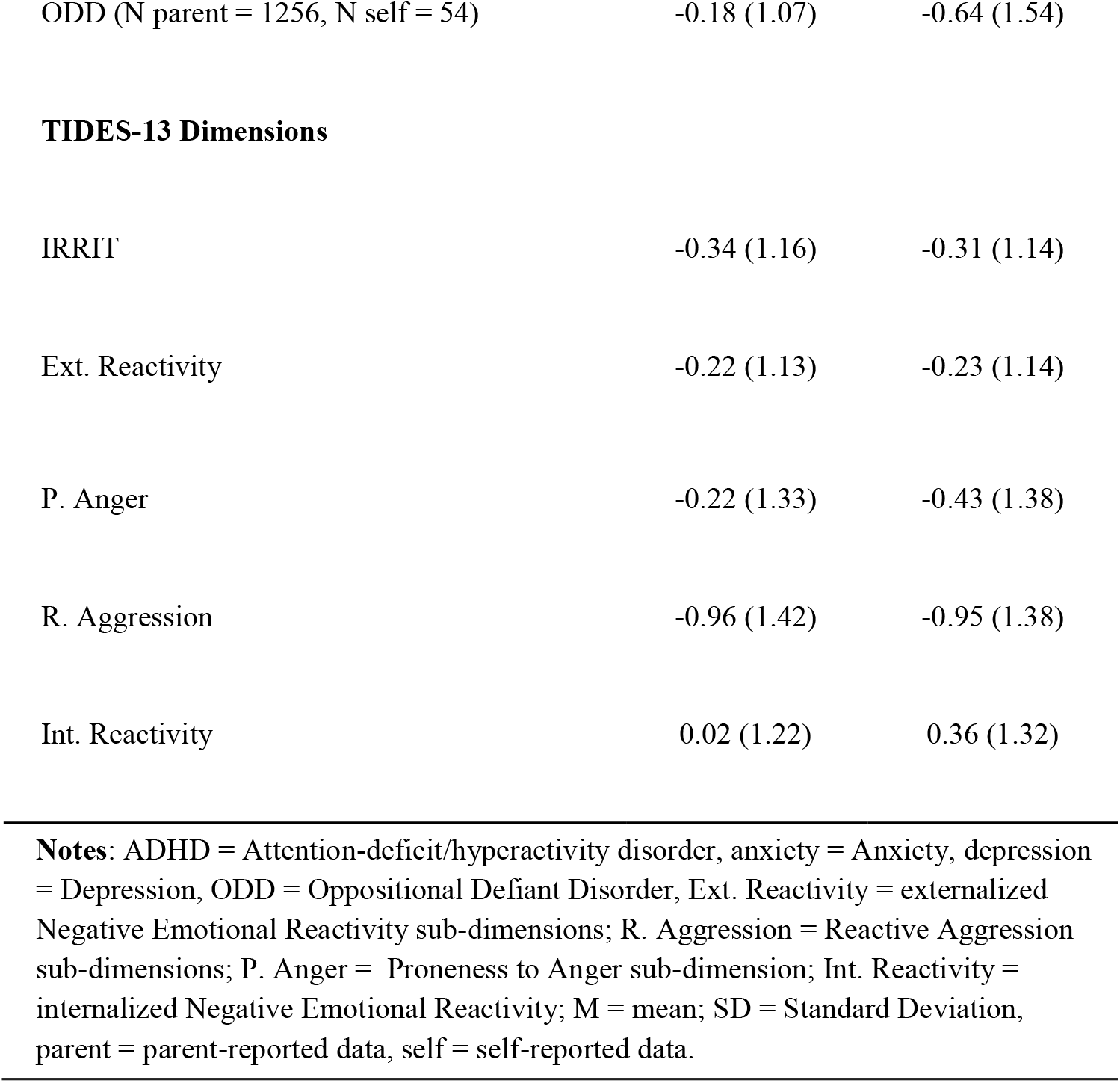
Sample Characteristics

### Ethical considerations

Informed consent, and verbal assent when applicable, were obtained from all participants, as approved by the local institution’s ethics review board.

### Predictive Validity

To assess the predictive validity of irritability sub-dimensions, a subsets of parents and youth completed the ARI (parent: N = 802; self: N = 77), as well as measures of ADHD (parent: N = 3867; self: N = 516) ODD (parent: N = 1256; self: N = 54), anxiety (parent: N = 1554, self: N = 109) and depression (parent: N = 1555; self: N = 109) traits. Anxiety and Depression traits were measured by summing the 6 item depression subscale and the 10 item subscale of the Revised Children’s Anxiety and Depression Scale-25 (RCADS).^23^ ADHD and ODD traits were measured by summing the 18 ADHD items and the 8 ODD items, respectively, in the Strength and Weakness of ADHD and Normal behavior scale (SWAN).^24^ SWAN scores were reversed so that higher scores reflected higher levels of the trait for consistency with the other measures.

### Statistical Analyses

To evaluate TIDES-13’s dimensionality, we randomly split the parent-report data into an exploratory factor analysis (EFA) sample (N = 1937) and a confirmatory factor analysis sample (CFA; N = 1938). The self-report sample (N = 516) was used entirely for CFA. Detailed procedure for the EFA can be found in the supplemental materials. In CFA, we used a robust maximum likelihood estimator (MLR) due to moderate violations to multivariate normality, as indicated by visual examinations of the Chi-Squared Q-Q plot. Model fit was assessed based on the following fit indices: Comparative Fit index (CFI;.95 and above indicate excellent fit whereas .90-0.95 indicate acceptable fit), the Tucker Lewis Index (TLI; values above .95 indicate excellent fit whereas .90-.95 indicate acceptable fit), the root means square error of approximation (RMSEA; values smaller than .08 indicate acceptable fit), and the standardized root mean square residual (SRMR; recommended cut-off at .08).^25^ We additionally tested the fit of multiple higher order models with a single higher order global dimension and the sub-dimensions extracted from EFA to assess the degree to which the sub-dimensions of irritability, in fact, contributed to a single underlying construct. To compare the relative contributions of a higher order global dimension versus each sub-dimension to the item’s variance, we performed Schmid-Leiman transformations^26^ to the best fitting higher order model, which enabled variance partitioning between global dimensions and the sub-dimensions. We also used the standardized loadings from the Schmid-Leiman transformations to calculate indices of internal consistency reliability (ie. the proportion of variance in the calculated score attributed to the construct of interest).^27^ We calculated omega total, omega hierarchical (ω_h_), and omega sub-dimension. Omega total estimates the proportion of variance of a total score attributable to all sources of common variance, where a high omega value reflects a highly reliable multidimensional composite.^28^ ω_h_ represents the proportion of variance attributable to a common factor and is a measure of general factor saturation,^28^ where a high ω_h_ indicates that the general factor is a dominant source of test score variance.^29^ Omega sub-dimension represents the internal consistency reliability of each sub-dimension score to measure the corresponding sub-dimension, over and above the global construct.^30^ Each omega value can range from 0 (no reliability) to 1 (perfect reliability). CFA models were fitted using lavaan^31^ and the Schmid-leimen transformation and omega values were calculated using EFAtools.^32^ Both packages are freely available in R Studio.

Multivariable regression models were fit to examine associations of TIDES-13 sub-dimensions with age and gender. Age by gender interactions were also examined, and a subsequent gender-stratified analysis was performed when the interaction term was significant. Parent and self-report data were combined to examine the associations between irritability and age along the full age continuum (ie. 4-19 yrs), with adjustment for respondent.

We evaluated the associations of each TIDES sub-dimension with anxiety, depression, ADHD and ODD traits by first calculating the Pearson correlations between each trait measure and each TIDES-13 sub-dimension in the subsample of participants with both measures available. We compared the TIDES-13 sub-dimensions correlation between other outcome variables and other sub-dimensions using Hitner’s, May and Silver’s method for comparing dependent correlations.^33^ We then fit multivariable regression models for each outcome with each TIDES sub-dimension as a predictor, adjusting for covariates such as age and gender. Parent-reported outcomes were predicted by parent-reported variables; self-reported outcomes were predicted by self-reported variables. To test the degree to which associations were driven by variance unique to specific sub-dimensions or rather by shared variance between each sub-dimension (i.e., the global factor), we performed a regression commonality analysis.^34^ Variance inflation factors (VIF) were also calculated to ensure the results were not severely degraded by multicollinearity. VIF’s under 10 were considered acceptable with no severe violations.^35^ Statistical significance was considered at *p* < .05 for all analyses.

## RESULTS

### Dimensionality

EFA and CFA revealed a four-dimension model with dimensions corresponding to Proneness to Anger (P. Anger), Reactive Aggression (R. Aggression), externalized negative emotional reactivity (Ext. Reactivity), and internalized negative emotional reactivity (Int. Reactivity) fit the data best in both parent-report and self-report. However, as model selection based fit statistics is generally unadvisable,^30^ we further examined the corresponding higher order model with the same four dimensions as sub-dimensions and an additional higher order global irritability (IRRIT) dimension (Figure 1) due to the moderate to high inter-dimension correlations (Table 2). This higher order model showed excellent fit in parent-report data (CFI_parent_ = .973, TLI_parent_ = .965, RMSEA_parent_ = .76, SRMR = .027) and self-report data (CFI_self_ = .971, TLI_self_ = .944, RMSEA_self_ = .78, SRMR = .044). The loadings of the higher order model with four sub-dimensions (Figure 1) revealed that each item influenced its corresponding sub-dimension (λ_parent_ > .74; Median λ_parent_ = .88; λ_self_ > .61; Median λ_self_ = .82), indicating that the sub-dimensions were well defined.

**Table 2:**
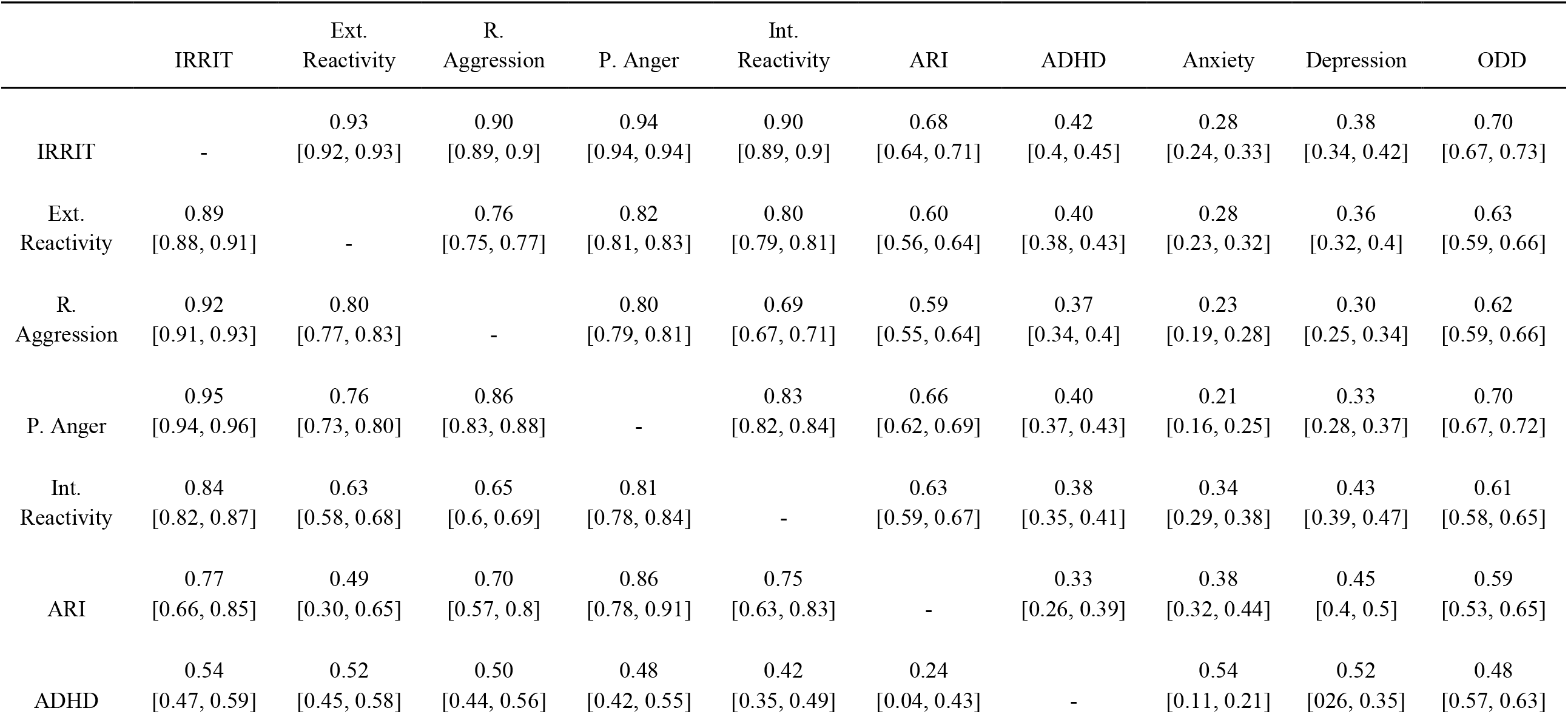
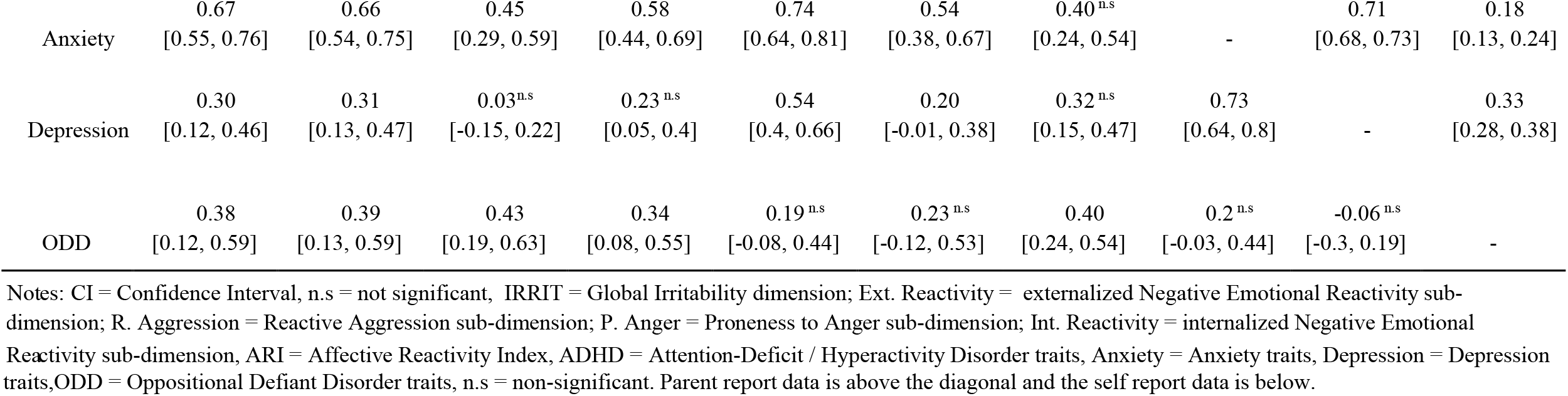
Pearson correlations and 95 CI between TIDES sub-dimensions, ARI and other psychopathology.

|  | IRRIT | Ext.<br>Reactivity | R.<br>Aggression | P. Anger | Int.<br>Reactivity | ARI | ADHD | Anxiety | Depression | ODD |
| --- | --- | --- | --- | --- | --- | --- | --- | --- | --- | --- |
| IRRIT | - | 0.93<br>[0.92, 0.93] | 0.90<br>[0.89, 0.9] | 0.94<br>[0.94, 0.94] | 0.90<br>[0.89, 0.9] | 0.68<br>[0.64, 0.71] | 0.42<br>[0.4, 0.45] | 0.28<br>[0.24, 0.33] | 0.38<br>[0.34, 0.42] | 0.70<br>[0.67, 0.73] |
| Ext.<br>Reactivity | 0.89<br>[0.88, 0.91] | - | 0.76<br>[0.75, 0.77] | 0.82<br>[0.81, 0.83] | 0.80<br>[0.79, 0.81] | 0.60<br>[0.56, 0.64] | 0.40<br>[0.38, 0.43] | 0.28<br>[0.23, 0.32] | 0.36<br>[0.32, 0.4] | 0.63<br>[0.59, 0.66] |
| R.<br>Aggression | 0.92<br>[0.91, 0.93] | 0.80<br>[0.77, 0.83] | - | 0.80<br>[0.79, 0.81] | 0.69<br>[0.67, 0.71] | 0.59<br>[0.55, 0.64] | 0.37<br>[0.34, 0.4] | 0.23<br>[0.19, 0.28] | 0.30<br>[0.25, 0.34] | 0.62<br>[0.59, 0.66] |
| P. Anger | 0.95<br>[0.94, 0.96] | 0.76<br>[0.73, 0.80] | 0.86<br>[0.83, 0.88] | - | 0.83<br>[0.82, 0.84] | 0.66<br>[0.62, 0.69] | 0.40<br>[0.37, 0.43] | 0.21<br>[0.16, 0.25] | 0.33<br>[0.28, 0.37] | 0.70<br>[0.67, 0.72] |
| Int.<br>Reactivity | 0.84<br>[0.82, 0.87] | 0.63<br>[0.58, 0.68] | 0.65<br>[0.6, 0.69] | 0.81<br>[0.78, 0.84] | - | 0.63<br>[0.59, 0.67] | 0.38<br>[0.35, 0.41] | 0.34<br>[0.29, 0.38] | 0.43<br>[0.39, 0.47] | 0.61<br>[0.58, 0.65] |
| ARI | 0.77<br>[0.66, 0.85] | 0.49<br>[0.30, 0.65] | 0.70<br>[0.57, 0.8] | 0.86<br>[0.78, 0.91] | 0.75<br>[0.63, 0.83] | - | 0.33<br>[0.26, 0.39] | 0.38<br>[0.32, 0.44] | 0.45<br>[0.4, 0.5] | 0.59<br>[0.53, 0.65] |
| ADHD | 0.54<br>[0.47, 0.59] | 0.52<br>[0.45, 0.58] | 0.50<br>[0.44, 0.56] | 0.48<br>[0.42, 0.55] | 0.42<br>[0.35, 0.49] | 0.24<br>[0.04, 0.43] | - | 0.54<br>[0.11, 0.21] | 0.52<br>[0.26, 0.35] | 0.48<br>[0.57, 0.63] |
| Anxiety | 0.67<br>[0.55, 0.76] | 0.66<br>[0.54, 0.75] | 0.45<br>[0.29, 0.59] | 0.58<br>[0.44, 0.69] | 0.74<br>[0.64, 0.81] | 0.54<br>[0.38, 0.67] | 0.40<sup>n.s</sup><br>[0.24, 0.54] | - | 0.71<br>[0.68, 0.73] | 0.18<br>[0.13, 0.24] |
| Depression | 0.30<br>[0.12, 0.46] | 0.31<br>[0.13, 0.47] | 0.03<sup>n.s</sup><br>[-0.15, 0.22] | 0.23<sup>n.s</sup><br>[0.05, 0.4] | 0.54<br>[0.4, 0.66] | 0.20<br>[-0.01, 0.38] | 0.32<sup>n.s</sup><br>[0.15, 0.47] | 0.73<br>[0.64, 0.8] | - | 0.33<br>[0.28, 0.38] |
| ODD | 0.38<br>[0.12, 0.59] | 0.39<br>[0.13, 0.59] | 0.43<br>[0.19, 0.63] | 0.34<br>[0.08, 0.55] | 0.19<sup>n.s</sup><br>[-0.08, 0.44] | 0.23<sup>n.s</sup><br>[-0.12, 0.53] | 0.40<br>[0.24, 0.54] | 0.2<sup>n.s</sup><br>[-0.03, 0.44] | -0.06<sup>n.s</sup><br>[-0.3, 0.19] | - |

Notes: CI = Confidence Interval, n.s = not significant, IRRIT = Global Irritability dimension; Ext. Reactivity = externalized Negative Emotional Reactivity sub-dimension; R. Aggression = Reactive Aggression sub-dimension; P. Anger = Proneness to Anger sub-dimension; Int. Reactivity = internalized Negative Emotional Reactivity sub-dimension, ARI = Affective Reactivity Index, ADHD = Attention-Deficit / Hyperactivity Disorder traits, Anxiety = Anxiety traits, Depression = Depression traits, ODD = Oppositional Defiant Disorder traits, n.s = non-significant. Parent report data is above the diagonal and the self report data is below.

**Figure 1:**
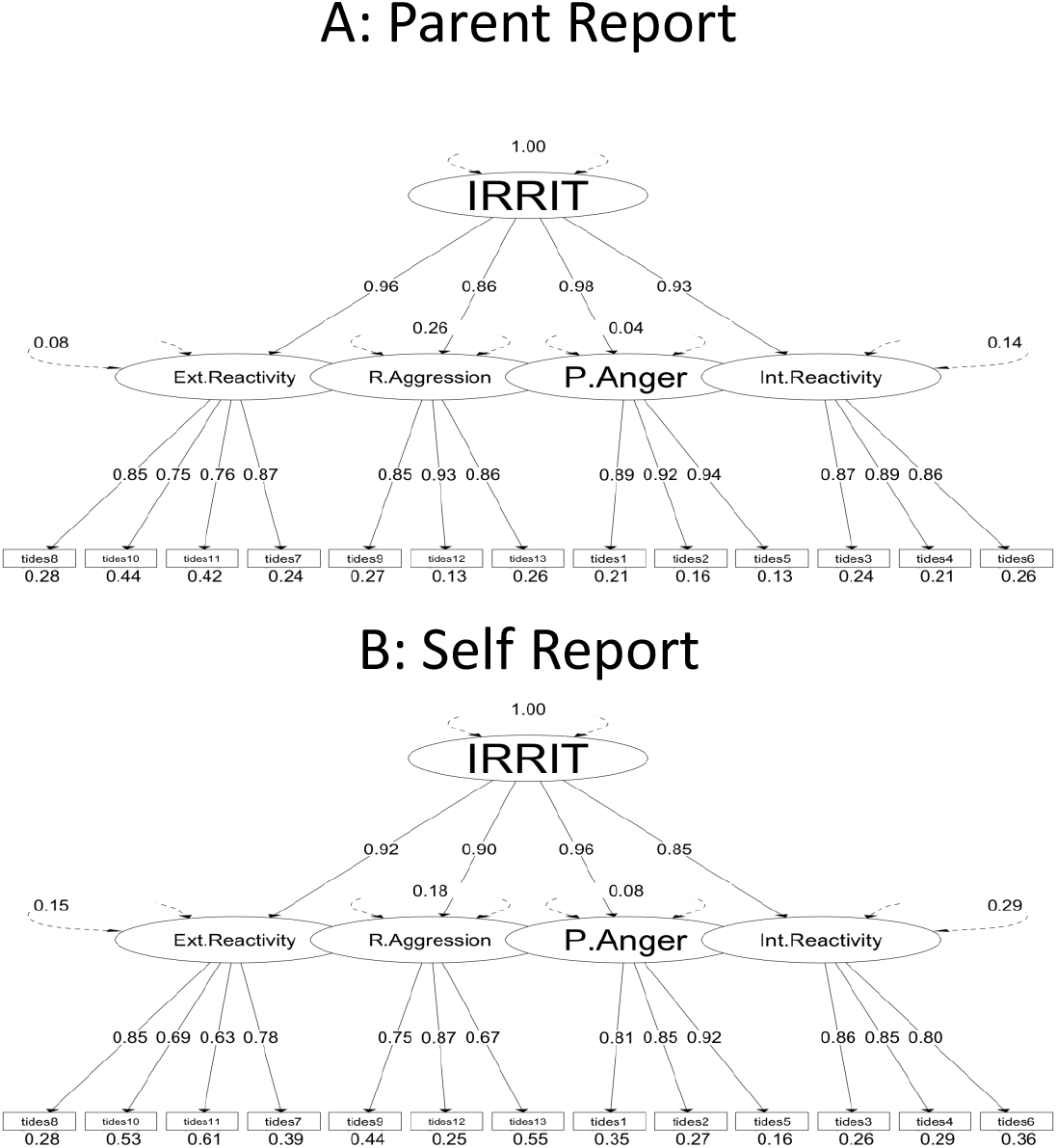
Higher Order Model of Irritability and Factor loadings. Both parent-reported (A) and self-reported (B) The Irritability and Dysregulation of Emotion Scale (TIDES-13) scores showed a higher order model of irritability with a global irritability dimension (IRRIT) and externalized negative emotional reactivity (Ext. Reactivity), reactive aggression (R. Aggression), proneness to anger (P. Anger), internalized negative emotional reactivity (Int. Reactivity). The numbers on the straight lines represent the standardized loading. The numbers places around the circular arrows represents the corresponding variable’s variance.

The loadings of the sub-dimensions onto the global irritability dimension (from λ_parent_ = .86 (R. Aggression) to λ_parent_ = .98 (P. Anger); from λ_self_ = .85 (R. Aggression) to λ_self_ = .96 (P. Anger) demonstrated that each the sub-dimension was strongly influenced by IRRIT (Figure 1). The high standardized loadings from the Schmid-Leiman transformation of the higher order four sub-dimension model (Table 3) also suggested a strong underlying global irritability dimension. The median of the standardized item loadings on IRRIT was λ = .81 (range: λ = .72 to λ = .92) in parent report and λ = .72 (range: λ = .58 to λ = .87) in self-report. However, each item also had consistent non-negligible loadings onto its corresponding sub-dimensions (Median λ_parent_ = .24; range: from λ_parent_ = .18 to λ_parent_ .46; Median λ_self_ = .24; range: from λ_self_ = .24 to λ_self_ = .46) which suggested that the sub-dimensions had incremental impact on the corresponding item scores over and above IRRIT. Descriptions of all models that were extracted in EFA and compared in CFA, along with their respective fit indices, can be found in the supplemental materials along with their respective fit indices in Table S1.

**Table 3:**
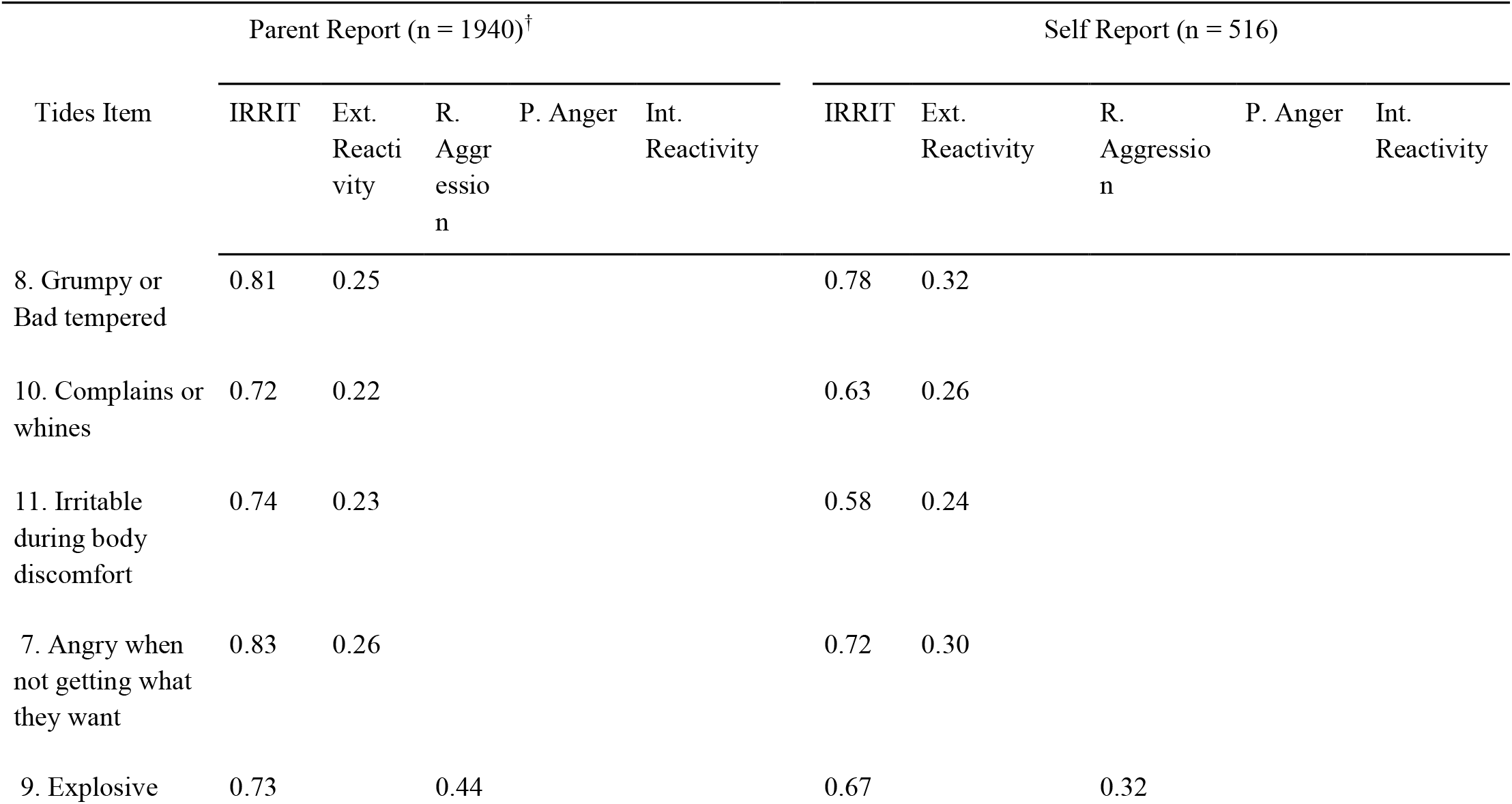
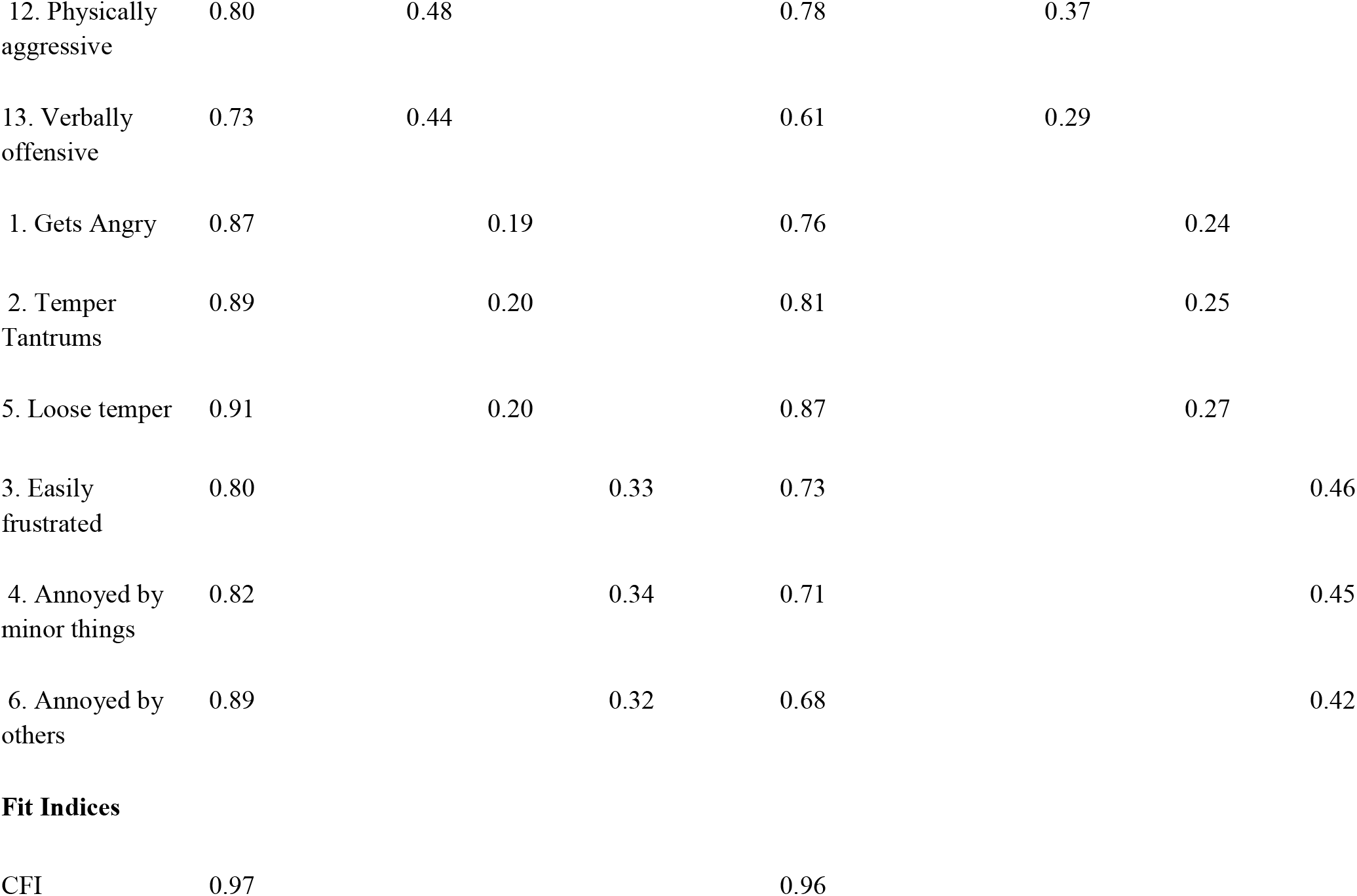
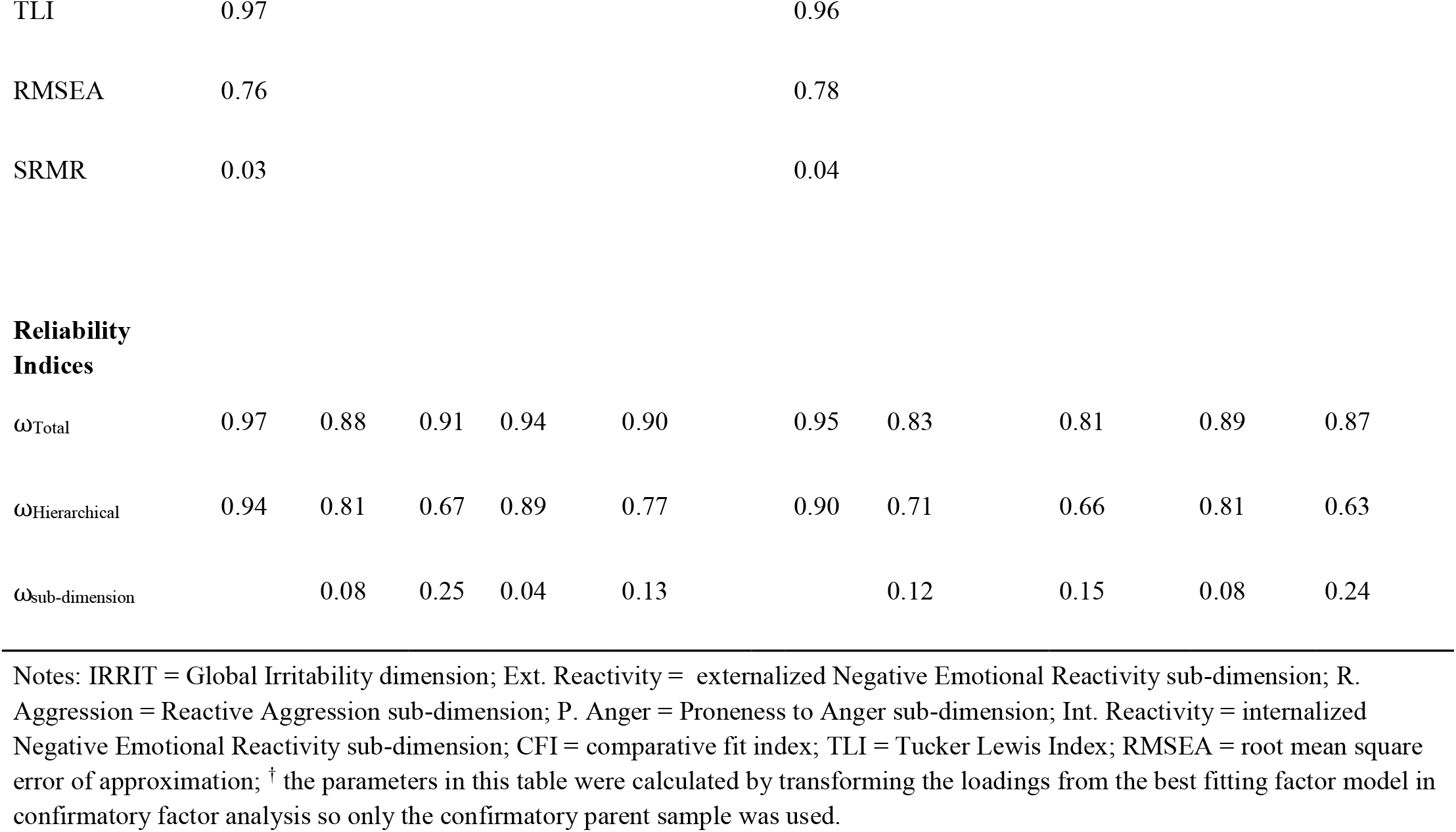
Schmid-Leiman Transformed Standardized factor loadings, Fit indices, and Reliability estimates of TIDES-13

Model-based omega estimates suggested that IRRIT had excellent internal consistency reliability and contributed to a large proportion of the total variance of all the items (Table 3). All estimated omega values can be found in Table 3. The ω_h_ values for each sub-dimension were satisfactory to excellent (Median ω_H parent_ = .81; range: from ω_H parent_ = .67 (R. Aggression) to ω _H parent_ .89 (P. Anger); Median ω_H self_ = .68; range: ω_H self_ = .63 to ω_H self_ = .81). The ω_h_ value corresponding to IRRIT was very high in parent report (ω_H_ = .94) and self report (ω_H_ = .90) suggesting a strong underlying global factor. Correspondingly, the omega sub-dimension values for the sub-dimensions were low (Median ω_parent_ = .11; range: ω_parent_ = .04 to ω_parent_ .25; Median ω_self_ = .14; range: ω_self_ = 0.08 to ω_self_ = .24), further confirming that a large proportion of the reliable variance was captured by IRRIT

### Predictive Validity of TIDES-13 sub-dimensions

In multivariable regression models examining age and gender effects, the age by gender interaction terms were significant for each sub-dimension. In subsequent gender stratified analyses, IRRIT, P. Anger, Ext. Reactivity and R. Aggression decreased with age in males but showed no significant associations with age in females whereas Int. Reactivity increased with age in females but showed no significant associations with age in males (Table S2).

In parent- and self-report, Int. Reactivity was significantly more correlated with anxiety traits than all the other TIDES-13 sub-dimensions (Table 2): P. Anger (parent: z = 6.47, p< .001; self: z = 3.53, p < .001), Ext. Reactivity (parent: z = 3.38, p < .001; self: z = 2.3, p = .021), and R. Aggression (parent: z = 5.58, p <0.001; self: z = 2.55, p = .011). Similarly, parent reported Int. Reactivity was also significantly more correlated with depression traits than all the other sub-dimensions (Table 2): P. Anger (z = 5.74, p < .001), Ext. Reactivity (z = 3.86, p < 0.001) and R. Aggression (z = 5.23, p < .001). Self-reported Int. Reactivity was significantly more correlated with depression traits than P. Anger (z = 2.64, p = .008) and R. Aggression (z = 2.37, p = .018) but no significant differences were found when compared to the correlation between depression traits and Ext. Reactivity (z = 1.69, p = .092).

As expected, P. Anger had the highest correlation with the ARI (Table 2). Parent-reported P. Anger was significantly more correlated with the ARI than Ext. Reactivity (z = 3.7, p <.001) and R. Aggression (z = 4.56, p < .001) however, no significant difference was found between the correlation of the ARI and P. Anger, and the correlation of the ARI and Int. Reactivity (z = 1.36, p = .174). Self-reported P. Anger was significantly more correlated with the ARI than Ext. Reactivity was (z = 1.20, p = .048), but no significant differences were found when comparing the correlation of the ARI and P. Anger to the correlation of the ARI and Int. Reactivity (z = 0.31, p = .757) and the correlation of the ARI and R. Aggression (z = 0.06, p = .952).

Parent-reported P. Anger was significantly more correlated with ODD traits than Int. Reactivity (z = 5.79), Ext. Reactivity (z = 3.2, p = .0013) and R. Aggression (z = 3.40, p = .0007). In self-report, although, R. Aggression had the greatest correlation with ODD traits, it was not significantly greater than the correlations between ODD traits and the other sub-dimensions

In multivariable regression models, Int. Reactivity was the strongest significant predictor (i.e., largest beta coefficient) of anxiety and depression (Table 4). A similar trend was seen in self-report, where Int. Reactivity was the sole significant predictor of anxiety and depression traits (Table 4). Further commonality analysis revealed that in parent report data, Int. Reactivity uniquely explained 31.1% and 23.9% of the total variance explained by the regression models predicting anxiety and depression traits respectively, whereas 41.5% and 54.4% of the variance explained was shared by all sub-dimensions. These results suggest IRRIT explains a substantial portion of the anxiety and depression trait variance, however, Int. Reactivity is uniquely associated with internalizing traits, over and above the associations of IRRIT. In self-report data, surprisingly, a greater proportion of the variance of anxiety and depression traits was uniquely explained by Int. Reactivity (77.8% and 78.3% respectively) than the variance explained by IRRIT (14.4% and 11.4% respectively), suggesting that Int. Reactivity primarily predicted internalizing traits, even more so than IRRIT.

**Table 4:**
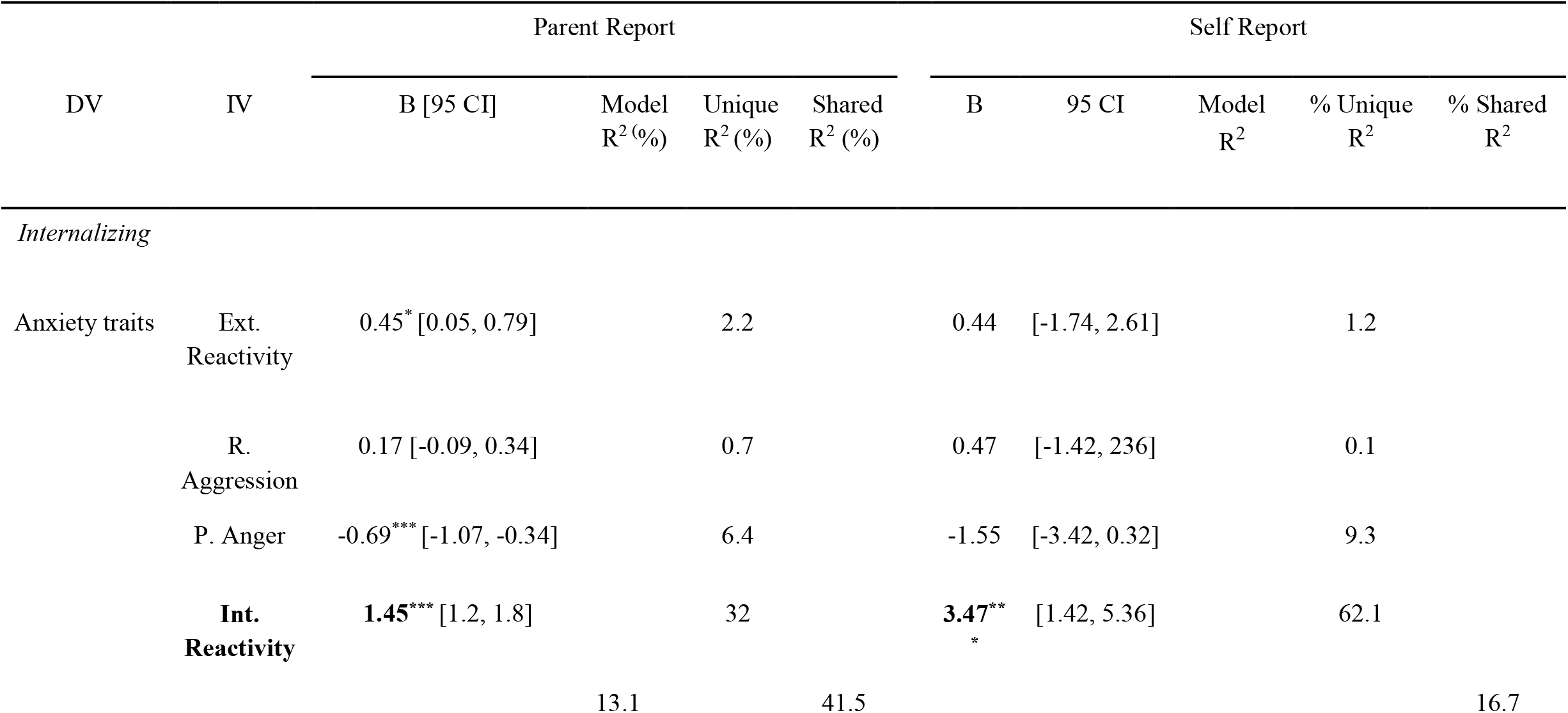
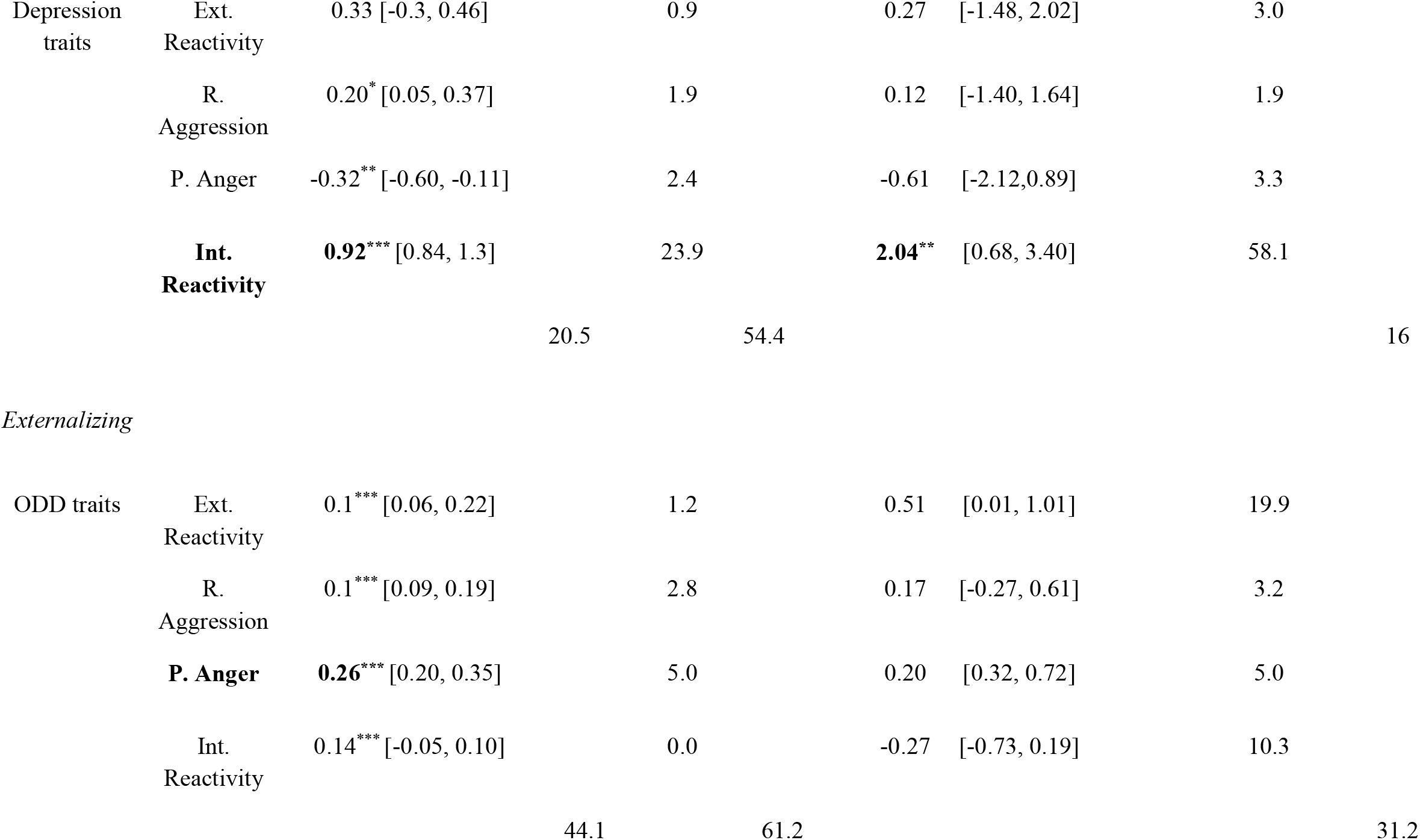
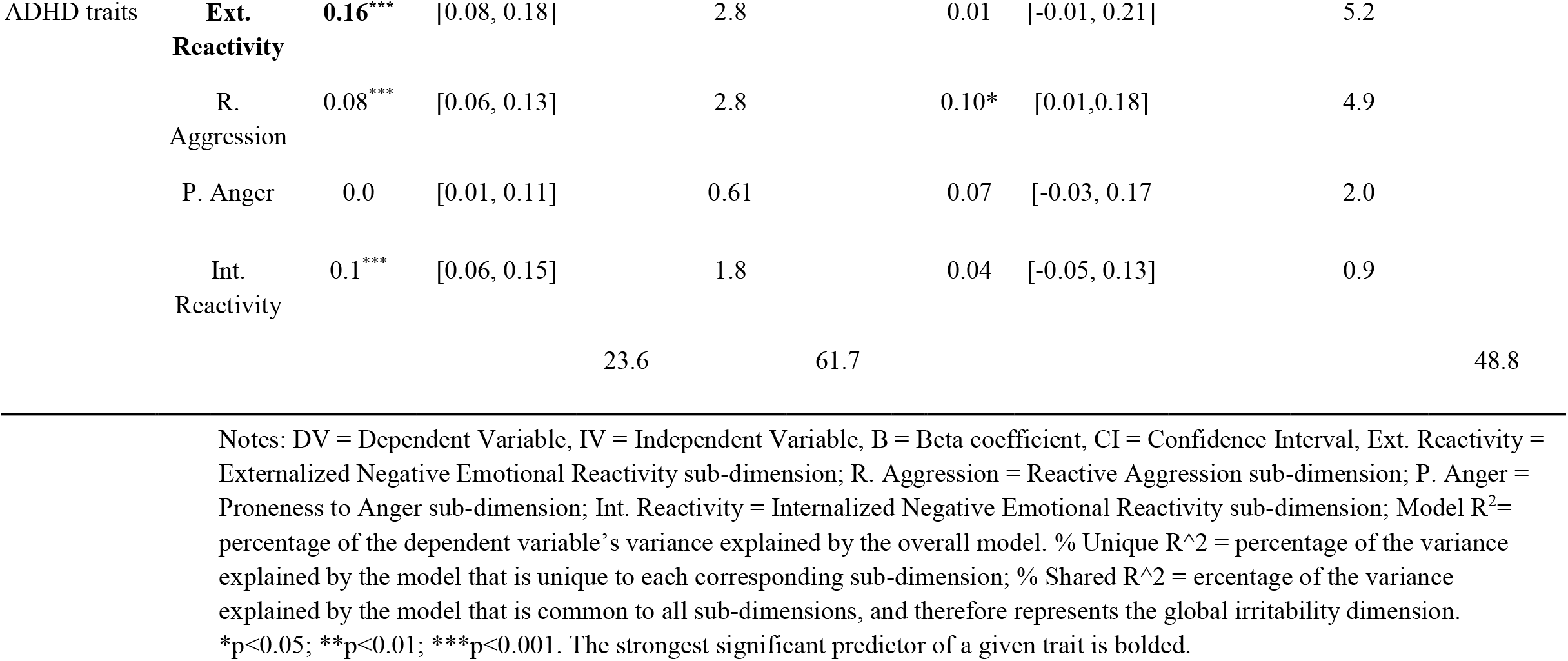
Multiple regression results of associations between TIDES sub-dimensions with Anxiety, Depression, ODD, and ADHD traits.

In multivariable regression models, parent reported P. Anger was the strongest predictor of ODD traits; however, substantially more of the variance explained by the model was shared by all sub-dimensions (60.4%) than what was uniquely explained by P. Anger (6.9%). In self-reported data, no sub-dimensions were significant predictors, however, there a limited number of self-report participants who had completed both the TIDES-13 and the ODD subscale (n = 54).

All sub-dimensions were similarly correlated with ADHD traits and had overlapping confidence intervals (Table 2). In regression models (Table 4), a majority of the variance explained was explained by IRRIT (parent report: % shared = 61.7, self report: % shared = 48.8).

## DISCUSSION

This article examined the heterogeneity of irritability symptoms in a large community sample using TIDES-13, a novel scale designed to capture various expressions of irritability. Our first aim was to use exploratory and confirmatory factor analyses to see if irritability sub-dimensions could be discerned. Our results lend further support to the notion that irritability is a heterogeneous construct with highly interrelated, yet distinct, ways of manifesting. We identified highly correlated, but distinguishable sub-dimensions that showed excellent fit across respondents (parent vs self): P. Anger, R. Aggression, Int. Reactivity, and Ext. Reactivity. Three of these sub-dimensions aligned with our hypothesis that dimensions relating to proneness to anger, reactive aggression, and internal affective state (Int. Reactivity) would emerge. However, within the remaining items, an external (Ext. Reactivity) response to stimuli also emerged.

The empirical distinction between P. Anger, characterized by anger outbursts, and Int. Reactivity, an internal persistent state, within our results aligns with the a priori differentiation between phasic (i.e., severe and developmentally inappropriate anger outbursts) and tonic (ie. persistent irritable mood between outbursts) irritability. The separation of these two sub-dimensions is also consistent with recent empirical findings that suggest phasic and tonic irritability are distinct, yet highly related irritability dimensions.^19,21^

The emergence of reactive aggression sub-dimension within our results is also important given the debate on whether notions of irritability should include behavioral aspects such as aggression. The result from our higher order modeling reveals that R. Aggression loaded onto the global factor (IRRIT) to a similar extent as other irritability sub-dimensions. This supports the conceptualization of R. Aggression as a distinguishable behavioral manifestation of irritability.

Given the calls for more nuanced irritability measures that are better able to differentiate irritability-related constructs such as tonic and phasic irritability from each other,^19^ having P. Anger, R. Aggression, Int. Reactivity, and Ext. Reactivity sub-dimensions within a measure of irritability may enable more nuanced explorations of the construct’s phenomenology. However, we also found a high degree of unity within the isolated sub-dimensions, which may explain why irritable sub-dimensions have been so difficult to distinguish previously. We observed that P. Anger, R. Aggression, Int. Reactivity, and Ext. Reactivity showed high degrees of interrelatedness and loaded strongly onto a global dimension (IRRIT) with high internal reliability. Furthermore, when evaluating the association of sub-dimensions with other forms of psychopathology, we saw that ADHD and ODD traits were driven primarily by IRRIT rather than individual sub-dimensions. We also observed high ω_h_ values of IRRIT (.94 and .90) suggesting that much of the item variance was driven by one underlying factor. These findings are consistent with studies conducted in a general population that suggest irritability sub-dimensions show high levels of inter-relatedness in normative samples.^13^ Paired with our findings, these studies suggest that within a normative sample, irritability can be considered essentially unidimensional implying the existence of a unique unidimensional latent trait in the presence of minor sub-dimensions.^36^ In light of these findings, we suggest that investigators using TIDES-13 to measure irritability in normative samples may utilize a total score as a valid and reliable measure of global irritability.

However, these results do not invalidate the potential of measuring irritability through sub-dimensions. Our results indicated that measuring irritability sub-dimensions provides additional information beyond irritability in general. We observed that Int. Reactivity increased with age in females and showed no association with age in males, contrary to the other sub-dimensions which decreased with age in males and showed no association in females. We also observed that Int. Reactivity was also uniquely associated with internalizing traits alluding to its discriminant validity. This differential association is consistent with work that suggests that mood-related irritability items predict depression in early adulthood over and above oppositional defiant dimension related items,^20^ lending support to the Int. Reactivity’s predictive validity. Given the emerging consensus that irritability in children is a longitudinal predictor of anxiety and depression,^3^ further examination of the longitudinal associations with TIDES-13 sub-dimensions may also yield a better understanding of whether these associations are primarily driven by specific sub-dimensions of irritability. Furthermore, recent evidence suggests that different subtypes of irritability with different developmental trajectories may exist.^22,37^ One of these isolated subtypes showed an increasing association with age and female preponderance similar to Int. Reactivity’s associations with age/gender, whereas other irritability subtypes show male preponderance and an inverse association with age, similar to the cross sectional trend observed in the remaining TIDES sub-dimensions.^22^ However, developmental studies often utilize overall irritability scores, combining items from the different potential sub-dimensions of irritability obscuring the possibility that these subtypes are driven by particular irritability sub-dimensions.

We also found that parent-reported P. Anger was more strongly correlated with the ARI than most other sub-dimensions confirming that ARI items tend to reflect proneness to anger. However, it should be noted that although the magnitude of correlation coefficients in self-report suggested a similar trend, many of the comparisons of correlation coefficients in self report were non-significant. This could be partially due to the limited power available due to the small subset of self respondents who completed both the ARI and TIDES-13 (N = 73). Also, in the parent report, the differences in correlation coefficients between P. Anger and the ARI and Int. Reactivity and the ARI were non-significant. This may be due to the ARI’s item content as five of the items relate to P. Anger, and one item taps into Int. Reactivity (i.e., “easily annoyed”). Further examination of the differences between P. Anger and Int. Reactivity may bring clarity as to whether any differences between the two constructs are being masked by combining into a single dimension within existing measures.

There are some limitations of this study that should be noted. The study sample was population-based, and thus the number of participants with mental-health diagnoses was proportional to that in a general population. However, an irritability scale with variable sub-dimensions such as TIDES-13 would enable evaluations of the factor structure in different clinical populations. Future validation studies for TIDES-13 should aim to replicate the factor structure and cross-sectional associations in clinical samples so that differences between irritability sub-dimensions can be further explored. The cross-sectional nature of our results also limited inferences about the stability and long-term predictive validity of irritability factor structure found in the current study. Given the available data, we were only able to assess TIDES-13’s internal reliability. Future studies should administer TIDES to multiple raters across different respondents and different time points to examine the scale’s test-retest and inter-rater reliability.

## CONCLUSIONS

In a large general population sample with both self and parent report measures, we found unity and diversity between irritability sub-dimensions. Irritability can manifest variably as highly interrelated sub-dimensions P. Anger, Int. Reactivity, Ext. Reactivity and R. Aggression with distinct correlates of psychopathology. These results support TIDES-13 as a reliable and valid measure which captures a broader spectrum of the construct’s phenotype and thus may be useful for research or clinical purposes. We specifically underscore a need to further examine the hypothesis that the association between irritability and internalizing psychopathology may be primarily driven by Int. Reactivity.

## Data Availability

All data produced in the present study are available upon reasonable request to the authors

## Acknowledgements

This work was supported by the Canadian Institutes of Health Research (R.J.S., MOP-93696 and P.D.A., MOP-106573) and the Alberta Innovates Translational Health Chair in Child and Youth Mental Health (P.D.A.). The statistical expert on the manuscript is Dr. Annie Dupuis.

## Supplementary material

**Table S1:** CFA Model Fit indices

| Model | Parent Report |  |  |  | Self Report |  |  |  |
| --- | --- | --- | --- | --- | --- | --- | --- | --- |
|  | CFI | TLI | RMSEA | SRMR | CFI | TLI | RMSEA | SRMR |
| Unidimensional | 0.902 | 0.899 | 0.139 | 0.047 | 0.883 | 0.859 | 0.125 | 0.058 |
| 2 Dimension | 0.947 | 0.935 | 0.1 | 0.033 | 0.9 | 0.878 | 0.116 | 0.054 |
| 3 Dimension | 0.964 | 0.954 | 0.087 | 0.031 | 0.939 | 0.921 | 0.096 | 0.054 |
| 4 Dimension | .979† | .972† | .068† | .023† | .966† | .955† | .070† | .040† |
| H 2 Dimension | 0.95 | 0.939 | 0.101 | 0.033 | 0.9 | 0.877 | 0.117 | 0.054 |
| H 3 Dimension | 0.962 | 0.952 | 0.088 | 0.030 | 0.929 | 0.911 | 0.099 | 0.051 |
| H 4 Dimension | 0.972 | 0.964 | 0.076 | 0.028 | 0.957 | 0.944 | 0.078 | 0.044 |

Notes: CFA = confirmatory factor analysis; CFI = comparative fit index; TLI = tucker lewis index; RMSEA = root mean square error of approximation; SRMR = standardised root mean residual; H = higher order model with additional higher order global dimensions, † = best fitting model. This table summarizes the confirmatory factor analysis fit indices of multiple factor models extracted from exploratory factor analysis conducted in a separate exploratory sample.

**Table S2:** Associations of TIDES sub-dimensions with Age and Gender

| *Dependent variable:* |  |  |  |  |  |  |  |  |  |  |  |  |  |  |  |
| --- | --- | --- | --- | --- | --- | --- | --- | --- | --- | --- | --- | --- | --- | --- | --- |
| Sample | IRRIT |  |  | P. Anger |  |  | Int. Reactivity |  |  | Ext. Reactivity |  |  | R. Aggression |  |  |
|  | Beta<br>(Std. Error) |  |  | Beta<br>(Std. Error) |  |  | Beta<br>(Std. Error) |  |  | Beta<br>(Std. Error) |  |  | Beta<br>(Std. Error) |  |  |
|  | Overall | Female | Male | Overall | Female | Male | Overall | Female | Male | Overall | Female | Male | Overall | Female | Male |
| Age | 0.015<br>(0.010) | 0.003<br>(0.011) | -<br>**0.028**<br>*<br>(0.011) | 0.007<br>(0.011) | -0.011<br>(0.012) | -<br>**0.045**<br>***<br>(0.012) | 0.045***<br>(0.010) | **0.033** **<br>(0.011) | 0.010<br>(0.012) | 0.009<br>(0.010) | -0.005<br>(0.011) | -<br>**0.041** ***<br>(0.011) | 0.002<br>(0.012) | -0.003<br>(0.013) | -<br>**0.050** ***<br>(0.013) |
| **Gender** |  |  |  |  |  |  |  |  |  |  |  |  |  |  |  |
| Female | ref. |  |  | ref. |  |  | ref. |  |  | ref. |  |  | ref. |  |  |
| Male | **0.676** ***<br>(0.117) |  |  | **0.733** ***<br>(0.129) |  |  | **0.045** ***<br>(0.124) |  |  | **0.641** ***<br>(0.117) |  |  | **0.814** ***<br>(0.142) |  |  |
| **Respondent** |  |  |  |  |  |  |  |  |  |  |  |  |  |  |  |
| Parent | ref. |  |  | ref. |  |  | ref. |  |  | ref. |  |  | ref. |  |  |
| Self | 0.080<br>(0.072) | **0.232** *<br>(0.095) | -<br>0.098<br>(0.108) | -0.082<br>(0.080) | 0.056<br>(0.106) | -<br>**0.245**<br>*<br>(0.120) | 0.199**<br>(0.076) | **0.360** ***<br>(0.100) | 0.010<br>(0.116) | 0.313***<br>(0.072) | **0.490** ***<br>(0.096) | 0.105<br>(0.108) | 0.172*<br>(0.087) | **0.234** *<br>(0.116) | 0.099<br>(0.132) |
| Age:Gender | **-0.055** ***<br>(0.012) |  |  | **-0.055** ***<br>(0.013) |  |  | **-0.048** ***<br>(0.012) |  |  | -<br>**0.063** ***<br>(0.012) |  |  | **-0.057** ***<br>(0.014) |  |  |

*Notes: Ext. Reactivity = Externalized Negative Emotional Reactivity sub-dimension; R. AGGR = Reactive Aggression sub-dimension; P. Anger = Proneness to Anger sub-dimension; Int. Reactivity = Internalized Negative Emotional Reactivity sub-dimension, Std. Error = standard error, ref = reference category for categorical variables. Beta coefficients estimated for categorical variables are in comparison to the reference category. \*p<0.05; \*\*p<0.01; \*\*\*p<0.001. This table summarizes the results from multiple regression analyses examining the association between TIDES-13 sub-dimensions with age and gender. Gender-stratified analyses were undertaken when the Age: Gender interaction term was significant.*

## TIDES-13 Development

We began by reviewing the existing definitions of irritability and identifying components within these definitions including proneness to anger (phasic irritability), negative emotional reactivity, grumpy or grouchy mood (tonic irritability), and reactive aggression. We then created a large pool of candidate items corresponding to these five components from existing irritability measures. Responses were measured from strengths (−3) to weakness (3) to capture the full distribution of irritability in a general population sample. The existing item pool was administered to a development sample consisting of parents of children attending a psychiatric clinic at [XXXXXXXXX] Hospital (n = 149). Parents responding to the items were asked a list of qualitative questions probing their interpretations of each item. These results informed item modification and deletion. Items were further deleted based on low item-scale correlation, low contributions to internal consistency, and considerations of face validity. After these final modifications, we arrived at the 13-item scale used in this present study. TIDES-13 has two parts: the first is 13-item measure of irritability traits on a continuum, where parents/youth were asked to rate the participant/themselves compared to others of the same age over the past year on a 7-point Likert scale ranging from -3 (far less than others) to 3 (far more than others). The second part of the scale contains four additional items measuring the frequency of temper outbursts, frequency of physically aggressive outbursts and level of the irritability-related impairment, all of which correspond to DSM-5 criterion for DMDD and IED to aid in establishing research diagnoses.

## Exploratory Factor Analysis (EFA) Methods

In EFA, the number of factors to retain was informed by parallel analyses and the scree test. EFA models were extracted using varimax rotations in anticipation of highly correlated factors. We assigned items to factors when the factor loading was greater than 0.4. All EFA analysis was conducted using the stats package that runs freely in R studio.

## Exploratory Factor Analysis Results

Parallel analysis suggested extracting four factors, however visual examination of the scree plot for “elbowing” was not clearly distinguishable between two, three and four-factor solutions so we extracted all one through four-factor solutions to determine the number of sub-dimensions that best described TIDES-13-dimensional structure. The first was a unidimensional model with all items loading onto one latent variable. The two-factor model had two factors: Reactive Aggression and the remaining items. The three-factor model had mood, phasic, and reactive aggression factors. The four-factor factor model had externalized negative emotional reactivity (Ext. Reactivity), reactive aggression (R. Aggr), proneness to anger (P. Anger), internalized negative emotional reactivity (Int. Reactivity) factors.

